# Precision Network Modeling of Transcranial Magnetic Stimulation Across Individuals Suggests Therapeutic Targets and Potential for Improvement

**DOI:** 10.1101/2024.08.15.24311994

**Authors:** Wendy Sun, Anne Billot, Jingnan Du, Xiangyu Wei, Rachel A. Lemley, Mohammad Daneshzand, Aapo Nummenmaa, Randy L. Buckner, Mark C. Eldaief

**Author notes:** Joint first author (Wendy Sun, Anne Billot). Joint senior author (Randy L. Buckner, Mark C. Eldaief). Correspondence (R.L.B), (M.C.E).

## Abstract

Higher-order cognitive and affective functions are supported by large-scale networks in the brain. Dysfunction in different networks is proposed to associate with distinct symptoms in neuropsychiatric disorders. However, the specific networks targeted by current clinical transcranial magnetic stimulation (TMS) approaches are unclear. While standard-of-care TMS relies on scalp-based landmarks, recent FDA-approved TMS protocols use individualized functional connectivity with the subgenual anterior cingulate cortex (sgACC) to optimize TMS targeting. Leveraging previous work on precision network estimation and recent advances in network-level TMS targeting, we demonstrate that clinical TMS approaches target different functional networks between individuals. Homotopic scalp positions (left F3 and right F4) target different networks within and across individuals, and right F4 generally favors a right-lateralized control network. We also modeled the impact of targeting the dorsolateral prefrontal cortex (dlPFC) zone anticorrelated with the sgACC and found that the individual-specific anticorrelated region variably targets a network coupled to reward circuitry. Combining individualized, precision network mapping and electric field (E-field) modeling, we further illustrate how modeling can be deployed to prospectively target distinct closely localized association networks in the dlPFC with meaningful spatial selectivity and E-field intensity and retrospectively assess network engagement. Critically, we demonstrate the feasibility and reliability of this approach in an independent cohort of participants (including those with Major Depressive Disorder) who underwent repeated sessions of TMS to distinct networks, with precise targeting derived from a low-burden single session of data. Lastly, our findings emphasize differences between selectivity and maximal intensity, highlighting the need to consider both metrics in precision TMS efforts.

---

Higher-order cognitive and affective functions are supported by distributed cortical association networks (Geschwind, 1965; Goldman-Rakic, 1988; Mesulam, 1990; 1998). Over the past few decades, human systems neuroscience approaches have yielded important insights into the organization and functional properties of association networks with recent focus on within-individual mapping (e.g., Fedorenko et al., 2010; Laumann et al., 2015; Braga and Buckner, 2017; Gordon et al., 2017; Somers et al., 2021). Leveraging the tools of precision neuroimaging to preserve idiosyncratic anatomical details in the individual, we recently reported on the intricate pattern of networks across the cortical mantle (and within prefrontal cortex) that subserve diverse higher-order cognitive and affective functions (Du et al., 2024). What was striking, and consistent with a growing number of reports (e.g., Mueller et al., 2013; Seitzman et al., 2019), is how variable functional anatomy is between individuals. This variability is unlikely methodological noise as individual estimates of network organization predict the topography of functional responses in independent task data. Thus, the between-individual differences arise from meaningful idiosyncratic differences in organization between people and therefore should be considered in precision approaches to neuromodulation.

Neuropsychiatric disorders have been associated with dysfunction in large-scale cortical association networks (Menon, 2011; Kaiser et al., 2015; Williams, 2016; Sha et al., 2019; Tozzi et al., 2021; Grot et al., 2024). Moreover, specific patterns of connectivity based on group-level network estimates have been variably linked to neuropsychiatric symptoms (Menon, 2011; Downar and Daskalakis, 2013; Xia et al., 2018); e.g., autonomic and interoceptive symptoms to the Salience network (Uddin, 2015), ruminative symptoms to the Default network (Chen et al., 2020; Zhou et al., 2020), and affect dysregulation symptoms to the Control network (Joormann and Vanderlind, 2014; Williams, 2016; Xia et al., 2018). Given the clinical heterogeneity of neuropsychiatric disorders, neuromodulation of symptom-specific networks may yield better therapeutic gains for certain indications (Eldaief et al., 2023; Siddiqi et al., 2020; Siddiqi and Fox, 2023). Importantly, traditional characterizations of functional networks have used estimates derived at the group level, averaging across many individuals to boost signal, which underrepresent idiosyncrasies observed in the individual patient (Cash et al., 2021; Moreno-Ortega et al., 2020). As such, targeting of one network based on group-level estimates may actually stimulate a different network in a given individual or multiple networks.

Transcranial Magnetic Stimulation (TMS) is an FDA-approved treatment for multiple neuropsychiatric disorders, including Major Depressive Disorder (MDD), Obsessive Compulsive Disorder (OCD), and migraines (e.g., Fox et al., 2012; Eldaief, Press, and Pascual-Leone, 2013; see Perera et al., 2016 and Trevizol et al., 2016 for reviews of TMS studies applied to MDD and OCD, respectively). Standard clinical targeting currently relies on anatomical landmarks, e.g., 4-5 cm anterior to the site where the motor threshold is obtained (Pascual-Leone et al., 1996); or modified EEG landmarks, e.g., left F3 and right F4 from the 10-20 coordinate system (Herwig, Satrapi, and Schönfeldt-Lecuona, 2003; Okamoto et al., 2004) or the Beam F3 method (Beam et al., 2009). As such, standard clinical approaches do not account for between-individual differences in functional network anatomy.

Increasingly, efforts have focused on establishing TMS targets based on functional neuroimaging (Hoffman et al., 2007; Mantovani et al., 2010; Eldaief et al., 2011; Fox et al., 2012; Wang et al., 2014; Santarnecchi et al., 2018; Nilakantan et al., 2019; Momi et al., 2020; Bagattini et al., 2021; Cash et al., 2021), as this has been hypothesized to improve clinical efficacy (Fox et al., 2012; Weigand et al., 2018). For instance, the accelerated Stanford Neuromodulation Therapy (SNT) TMS protocol employs a personalized targeting strategy. The SNT protocol, which was recently approved by the FDA for the treatment of MDD (Cole et al., 2020; 2022), targets a region of dorsolateral prefrontal cortex (dlPFC) anticorrelated to the subgenual cingulate cortex (sgACC). A recent study suggests that SNT alters connectivity within a group-level atlas-based estimate of the distributed network commonly referred to as the Default network (Gajawelli et al., 2024). An open question is how networks defined within individuals relate to the precision targeting approach deployed in the SNT protocol. That is, does defining targets based on anticorrelation with the sgACC consistently target a specific association network and, if so, which one?

We recently mapped and validated a precision network estimation approach using a novel Multi-Session Hierarchical Bayesian Model (MS-HBM) in 15 healthy individuals (Du et al., 2024). Here, we leverage this work and recent advances in precision network-level TMS targeting (Lynch et al., 2022) to evaluate which networks, defined at the individual level with precision functional MRI (fMRI), are stimulated across individuals when two of the available clinical protocols are used: (1) scalp-based landmarking (e.g., left F3 and right F4) and (2) sgACC anticorrelation targeting (e.g., as used in the SNT protocol). Further, we integrated a protocol for characterizing, at the individual level, the spatial selectivity and E-field intensity of personalized TMS network targets in the dlPFC. To do so, we combined tools from individualized preprocessing, precision network mapping, and electric field (E-field) modeling in an automated pipeline aimed to guide clinical neurostimulation decisions. The precision TMS pipeline was first implemented in a cohort of 15 intensively sampled healthy participants to gain insights on the functional network impact of existing clinical TMS protocols and demonstrate the potential for network-specific TMS targeting. The pipeline was then applied on an independent cohort of participants (including those with MDD) with fMRI data acquired in a single session to prospectively target distinct networks and retrospectively estimate network engagement. The impact of this work lies in its potential to tailor neurostimulation to individual brain networks, assess the network-level effects of TMS protocols, and improve functional precision of neuromodulation in both research and therapeutic settings.

## Materials and Methods

### Study Design

This study used neuroimaging data from two cohorts of participants. The first cohort included 15 intensively sampled participants enrolled as part of a broader study of network topography and functional mapping within the individual (Du et al., 2024; Kosakowski et al., 2024). Participants completed scanning across 8-11 sessions. For each participant, networks were estimated within the individual and the results run through the openly available Targeted Functional Network Stimulation (TANS) protocol for E-field modeling and target optimization (Lynch et al., 2022). Pertinent visualizations and results were automatically compiled into a comprehensive TMS planning report for each participant. From these precision modeling results, we gleaned insights into the networks impacted by common clinical TMS strategies as well as the potential to distinctly target side-by-side networks. The same precision TMS modeling approach was applied on an independent cohort of 8 participants, including 4 participants with MDD. These participants completed one scanning session (less than one hour of fMRI data collected) and received repetitive TMS (rTMS; see *TMS protocol*) to the modeled sites.

### Participants

A first cohort of 15 healthy right-handed adults aged 18-34 yr (mean = 22.1 yr, SD = 3.9 yr, 9 female), and a second cohort of 4 healthy adults aged 26-45 yr (mean = 34.8 yr, SD = 7.8 yr, 1 female) and 4 adults with MDD aged 24-39 yr (mean = 33.0 yr, SD = 7.0 yr, 2 female), representing diverse racial and ethnic backgrounds (12 non-White or Hispanic in total), participated for payment. The first cohort of participants are labeled *P1-P15*. Participants provided informed consent using a protocol approved by the Institutional Review Board at Harvard University (first cohort of participants) and Mass General Brigham (second cohort of participants).

### MRI Data Acquisition

For the jirst cohort, details of the methods have been reported previously (Du et al., 2024) and relevant aspects are repeated here. For the second cohort, details of the methods were largely the same, with minor differences noted below. All neuroimaging data were collected at the Harvard Center for Brain Science using a 3-T Prisma-fit MRI scanner and 32-channel head coil (Siemens Healthineers, Erlangen, Germany). Participants were monitored closely for motion using Framewise Integrated Real-time MRI Monitoring (FIRMM; Dosenbach et al., 2017) and for alertness using the EyeLink 1000 Core Plus with Long-Range Mount (SR Research, Ottawa, ON, Canada).

fMRI data were acquired with a custom multiband gradient-echo echo-planar pulse sequence provided by the University of Minnesota sensitive to blood oxygenation level-dependent (BOLD) contrast (e.g., Feinberg et al., 2010; Xu et al., 2013): voxel size = 2.4 mm, TR = 1,000 ms, TE = 33.0 ms, flip-angle = 64°, matrix 92 x 92 x 65 (FOV = 221 x 221), 65 slices, anterior-to-posterior (AP) phase encoding, multislice 5x acceleration. 17-24 resting-state fixation runs were collected for each participant, during which they fixated on a central black crosshair. Each run was 7 min 2 s, with 422 frames; the first 12 frames were removed for T1 equilibration. To mitigate spatial distortions, dual-gradient-echo B0 field maps were acquired at each session (TE=4.45, 6.91ms with slices matched to the BOLD sequence).

For *P1-P15* (first cohort), high-resolution T1-weighted (T1w) and T2-weighted (T2w) structural images were acquired based on the Human Connectome Project (HCP) sequences (Harms et al., 2018). T1w MPRAGE parameters: voxel size = 0.8 mm, TR = 2,500 ms, TE = 1.81, 3.60, 5.39, and 7.18 ms, TI = 1,000 ms, flip angle = 8°, matrix 300 x 320 x 208, 208 slices, in-plane GRAPPA acceleration = 2. T2w SPACE parameters: voxel size = 0.8 mm, TR = 3,200 ms, TE = 564 ms, matrix = 300 x 320 x 208, 208 slices, in-plane GRAPPA acceleration = 2. As backup, rapid T1w structural scans were also collected using a multi-echo MPRAGE sequence (van der Kouwe et al., 2008): voxel size = 1.2 mm, TR = 2,200 ms, TE = 1.57, 3.39, 5.21, and 7.03 ms, TI = 1,100 ms, flip angle = 7°, matrix 192 x 192 x 144, 144 slices, in-plane GRAPPA acceleration = 4.

For the second cohort, high-resolution T1-weighted (T1w) and T2-weighted (T2w) structural images were acquired using FreeSurfer sequences. T1w MPRAGE parameters: voxel size = 1.0 mm, TR = 2,530 ms, TE = 1.69, 3.55, 5.41, and 7.27 ms, TI = 1,100 ms, flip angle = 7°, matrix 256 x 256 x 192, 192 slices, in-plane GRAPPA acceleration = 2. T2w SPACE parameters: voxel size = 1.0 mm, TR = 3,200 ms, TE = 564 ms, matrix = 256 x 256 x 192, 192 slices, in-plane GRAPPA acceleration = 2. Backup T1w structural scans were collected using a multi-echo MPRAGE sequence with the same parameters as for the first cohort. One participant in the second cohort had the same T1w and T2w sequence as used for the first cohort, and another participant had the same T2w sequence as used for the first cohort. Due to artifacts in acquisition for one participant, the backup 1.2mm T1w image was used for preprocessing and modeling.

### Data Quality Control

Raw images were uploaded into an instance of the eXtensible Neuroimaging Archive Toolkit (XNAT; Marcus et al., 2007), an open-source data management and quality control (QC) platform. Automated QC was run including motion traces, slope images of drift, estimates of mean and absolute motion, and calculation of voxel and slice-based signal-to-noise ratio (SNR). MRI images were manually checked for incomplete head coverage, ringing, and ghosting. Exclusion criteria for resting-state fixation runs included: maximum absolute motion >1.8 mm and slice SNR <130. In the first cohort, on a case-by-case basis, runs with SNR <130 but >100 were inspected and kept if motion traces and visual inspection showed adequate quality. In the second cohort, all included runs had SNR >130, and one borderline rest run was included with motion of 1.86 mm. Data exclusions were finalized before any further analysis, and there were a total of 15-24 usable resting-state fixation runs (105-169 min of data) in each participant in the first cohort, and 5-8 runs (35-56 min of data) in each participant in the second cohort.

### Automated Network Mapping and TMS Targeting Pipeline in the Individual

An automated pipeline that integrates each step from MRI preprocessing to TMS planning was constructed and is made available as an open-source toolkit. The toolkit uses openly available software packages for MRI data processing and TMS E-field modeling, The steps in the processing pipeline were as follows.

*(1) MRI Preprocessing*. Raw neuroimaging data were preprocessed with an openly available pipeline (“iProc”), which minimizes spatial blur and preserves individual idiosyncratic anatomy with a single interpolation step for BOLD data (see detailed description in Braga et al., 2019), using tools from FreeSurfer (Fischl, 2012), FSL (Jenkinson et al., 2012), and AFNI (Cox, 2012). For the first cohort, preprocessed data were taken directly from Du et al. (2024). For the second cohort, data were preprocessed as follows: high-resolution T1w and T2w images were used for pial and white matter boundary estimation with FreeSurfer *recon-all*. Brain extraction was performed using FSL *BET*. A within-individual mean BOLD template was calculated by taking the average of the field map-unwarped, upsampled to 1.2 mm middle volumes of all runs that were registered to an unwarped, upsampled middle volume from a single run. For each BOLD run, 4 matrices were calculated to 1) align all volumes in a run to the middle volume of that run (FSL *FLIRT*, 12 Degrees of Freedom; DOF), 2) correct for geometric distortions caused by susceptibility gradients using a session-specific B0 field map to unwarp (FSL *FUGUE*), 3) register the unwarped BOLD volume to the within-subject mean BOLD template (FSL *FLIRT*, 12 DOF), and 4) register data from the mean BOLD template to the T1w native-space template, resampled to 1.0 mm (FreeSurfer *bbregister*, 6 DOF). These 4 matrices were composed into a single transform that was applied to raw BOLD volumes in a single interpolation to reduce spatial blur. A QC dashboard with the resampled T1w and interpolated BOLD images was generated, allowing for extensive checking for registration errors through web-based viewing and capture in an encapsulated PDF report.

After the single interpolation step, confounding variables including 6 head motion parameters, whole-brain signal, ventricular signal, deep cerebral white matter signal, and temporal derivatives as well as the quadratic term were calculated from the data (36 parameters; Ciric et al., 2017). In addition, volumes with high framewise displacement (defined as >0.4mm or >3 standard deviations above the mean) were flagged in each run and added to the 36-parameter nuisance regression matrix. These signals were regressed out (AFNI *3dTproject*), then data were bandpass filtered at 0.01–0.1-Hz (AFNI *3dBandpass*), projected to the fsaverage6 cortical surface mesh using trilinear interpolation, and then smoothed using a 2-mm full-width at half-maximum Gaussian kernel along the cortical surface (FreeSurfer *mri_vol2surf* and *mri_surf2surf*).

*(2) Within-Individual Cortical Network Estimation*. Within-individual cortical networks were estimated with a 15-network MS-HBM (see Kong et al., 2019; Du et al., 2024). For this study, network estimates were used directly from Du et al. (2024), and the steps are summarized here.

Using the available resting-state fixation runs for each individual as input, the MS-HBM allocated all fsaverage6 cortical surface vertices to one of 15 networks. For each run, the model calculated correlations between the time series at each of the 40,962 surface vertices and 1,175 regions of interest uniformly spread across the surface. An initial functional connectivity profile was defined as a binarized map of the top 10% of these correlations. The MS-HBM was then initialized with a 15-network group-level prior derived from the HCP S900 data release and used parameters derived from an expectation-maximization algorithm to estimate network assignments from the functional connectivity profiles of the individuals to be mapped. The MS-HBM was run in groups of 5 participants to yield network assignments for all 15 participants. A dashboard / PDF of 8 views including frontal, lateral, parietal, and medial, in both hemispheres was generated for QC and visualization.

Critically, to ensure that network estimates were not overly driven by model assumptions, a model-free seed region-based control check was performed (see Du et al., 2024). Seed region correlation utilized a Fisher *r*-to-*z* transformed 81,924 x 81,924 correlation matrix composed from all pairwise correlations between the fMRI time courses at each surface vertex during each resting-state fixation run, averaging across all runs. In addition, cortical network estimates derived from the MS-HBM showed high reliability across sessions (on average > 80% correspondence of networks estimated using independent data sets within individuals), as demonstrated in Fig. 2-4 of Du et al. (2024). The correspondence of network estimates improved with longer scanning times (see Supplemental Fig. S14 of Du et al., 2024).

The surface-based network estimates were projected back to the native-space surface using a series of Connectome Workbench (Marcus et al., 2011; v1.3.2) commands. First, the spherical registration file for each hemisphere (e.g., lh.sphere.reg) from FreeSurfer was converted to a Geometry Interchange Format (GIFTI) surface file with *wb_shortcuts -freesurfer-resample-prep* (e.g., lh.sphere.reg.surf.gii). Next, using the spherical registration GIFTI file, network estimates in each hemisphere were resampled to a high-resolution native space surface file with the “*ADAP_BARY_AREA*” method in *wb_command -label-resample*. With this method, native-space network estimates in each hemisphere were then resampled to the fsLR-32k space (FreeSurfer, left-right-symmetric, ∼32k vertices; Van Essen et al., 2012), which preserves individual cortical details while reducing computational burden with 32,492 vertices per hemisphere.

*(3) dlPFC Search Space*. A custom dlPFC search space, wherein individualized stimulation targets were ultimately defined, was created separately in both hemispheres. Starting from published atlas coordinates for the approximate location of Brodmann Area 46 (Fox et al., 2012; Rajkowska and Goldman-Rakic, 1995), a 30mm-radius spherical region of interest (ROI) centered at (-44, 40, 29) in Montreal Neurological Institute (MNI) space was created on the left, as well as its homolog on the right, centered at (44, 40, 29). The sphere was then shifted dorsally to allow for targeting of more networks, with a final center of (-34, 40, 36) on the left and (34, 40, 36) on the right. These ROIs were projected to the surface, regions in the insula and on the midline were removed, and final ROIs were resampled to fsLR-32k space for use in each individual. See **Fig. 4B** for the left dlPFC search space in one example participant. Note that this pipeline can be used with other search spaces.

*(4) E-field Modeling of Precision Targeting.* The TANS pipeline developed by Lynch, Liston and colleagues (Lynch et al., 2022) was adapted and applied to derive optimal coil positions and orientations to target each network set. Prior to TANS, all individual surface data, including network estimates, search spaces, pial and white matter surfaces, and sulcal depth maps were resampled to fsLR-32k space (Van Essen et al., 2012).

To begin, a tetrahedral head model including tissue segmentations was created using each participant’s T1w structural image (SimNIBS *charm*; Puonti et al., 2020). Next, a target region was identified within the left dlPFC search space. The target network(s) were masked within the search space, and vertices in the sulci were removed (thresholded at 0 using the .sulc files created by FreeSurfer *recon-all*) so that only the gyral crowns remained. The surface area of each remaining contiguous cluster was calculated, and the final target region was selected as the cluster with the largest surface area. After the target region was identified, a search grid was constructed on the scalp above the target region centroid. This search grid was then uniformly subsampled.

At each point in the subsampled search grid, an E-field simulation was run with SimNIBS (Thielscher, Antunes, and Saturnino, 2015) using a standardized stimulation intensity (1 A/μs) and coil to scalp distance (2 mm), with a MagVenture Cool-B70 coil model (MagVenture, Inc., Farum, Denmark). At this step, the standardized stimulation intensity from TANS was used, as E-field strength varies linearly with stimulation intensity; thus, it would not affect the optimal coil position (see Supplemental Fig. S2C of Lynch et al., 2022). The optimal coil position was selected from these simulations, based on an adapted form of the “on-target value” specified in Lynch et al. (2022). In brief, the on-target value was defined as the surface area of the target network(s) in the E-field thresholded at 99.0-99.9% (top 0.1-1% of non-zero values averaged across thresholds) divided by the total surface area of the thresholded E-field. At the optimal coil position, angles at 5° increments were tested, and the optimal orientation angle was identified to further maximize the “on-target value”. The final coil position (represented as the x,y,z coordinates where the coil center should be) and orientation (represented as the x,y,z coordinates toward which the coil handle should point) were saved. Finally, to characterize optimal “dose,” a range of stimulation intensities was tested at the optimal coil placement. Assuming a neural activation threshold of 100 V/m, suprathreshold E-field values were quantified for each network at each intensity level, and the level with the highest on-target value was recorded.

*(5) Spatial Selectivity.* For each E-field map, the spatial selectivity for networks was calculated at various thresholds ranging from 99.0-99.9% (0.1% increments). For example, at threshold 99.5%, the top 0.5% of the non-zero E-field values are kept, and the network assignment for each vertex in this thresholded map is pooled into a bar plot representing the % of vertices in the E-field map that belong to each network. For example selectivity plots, see **Fig. 1C**. Each row in the plot represents a different threshold, and each color represents a different network. This selectivity measure is adapted from Lynch et al. (2022).

*(6) E-field Intensity.* One novel contribution of this effort is a systematic measure of E-field intensity supplied to networks. This was calculated for each non-thresholded E-field map by applying an empirical cumulative density function (using R *ecdf*) to the E-field values and quantifying the intensity distribution within each network. This measure was motivated by work demonstrating high variability in TMS-induced neuronal activation in nearby neurons with maximal stimulation directly under the coil (Romero et al., 2019) and a positive relationship between electric field strength and the likelihood of TMS-induced evoked potentials (Wang et al., 2024).

**Fig. 1D** shows example intensity plots. Each row in the plot represents a different network. In this study, the E-field was calibrated to 120% of the average motor threshold (MT) for our center’s studies (mean MT = 29.2%, SD = 5.9% Machine Standard Output; MSO). Thus, the E-field was calibrated to 35% MSO, dI/dt = 48 A/μs for a MagVenture Cool-B70 coil.

The following sections describe how the steps above were applied to characterize, in terms of selectivity and intensity, the networks targeted by scalp-based landmarking (left F3 and right F4) and by sgACC anticorrelations, as well as to prospectively target side-by-side association networks in dlPFC.

### Left F3 and Right F4

To estimate network engagement at the scalp landmarks used in current clinical TMS protocols, the “eeg_positions” output from the SimNIBS *charm* function, left F3 and right F4 coil positions, were estimated in each participant based on the EEG 10-20 Okamoto coordinate system (Okamoto et al., 2004; Thielscher, Antunes, and Saturnino, 2015). SimNIBS v4.0.1 coil orientations were calculated by rotating the position 45° along the sagittal plane (Janssen, Oostendorp, and Stegeman, 2015) to approximate what is used in the clinic. E-field simulations were performed using these coil position / orientation vectors (step 4). Network engagement at left F3 and right F4 was then characterized with the precision TMS pipeline (steps 5 and 6).

### Within-Individual sgACC Correlations

Within each participant, a bilateral sgACC ROI was constructed using published coordinates in MNI space (Fox et al., 2012). The ROI, defined as a 10 mm radius sphere centered at (6, 16, -10), was reflected along the sagittal plane to yield a bilateral ROI, and projected to the fsaverage6 surface (Wu et al., 2018). The ROI was then masked to exclude vertices with SNR < 20 using individual-specific SNR maps (Du et al., 2024), as the sgACC falls within a region highly vulnerable to susceptibility artifacts (Ojemann et al., 1997). This ROI was used to generate an sgACC functional connectivity map of the cerebral cortex for each individual. Pearson’s correlations between the fMRI time series at the sgACC ROI and each cortical surface vertex were computed for each fMRI run, resulting in a 1 x 81,924 matrix (40,962 vertices per hemisphere). The matrices were *r*-to-*z* transformed and averaged across runs to yield a mean matrix with high stability. These maps were resampled to fsLR-32k space and used to identify regions that were negatively (*z*(r)<0) correlated to the sgACC. Then, an anticorrelation threshold was defined as the top 40% of negatively correlated voxels in dlPFC. Thresholded sgACC anticorrelations in example participants are shown in the first row of **Fig. S2**. The anticorrelated regions were used in the E-field modeling of precision targeting step of the precision TMS pipeline (step 4), and characterization of network engagement was performed (steps 5 and 6).

Critically, multiple methods and thresholds were tested, with little impact on results. For example, a liberal approach was tested whereby all negatively correlated regions in dlPFC were considered anticorrelated (*z*(r) < 0), as well as a strict variant of this approach in which only the top 100 vertices were selected as the anticorrelated target region (**Fig. S3**). Both methods yielded similar modeling outcomes as the 40% method. In addition, several thresholds – 20%, 40%, and 60% – were tested for defining anticorrelations, and this also did not markedly change the pattern of results (**Fig. S4**). That is, the results presented are robust to a range of parameter choices.

### Prospective Network Targeting

The precision TMS pipeline was applied to prospectively target individual-specijic side-by-side networks in dlPFC. In this study, we modeled the targeting of jive association networks, grouped into three distinct sets: Default Network-A & Default Network-B (DN-A & DN-B), Salience & Cingulo-Opercular (SAL & CG-OP) Networks, and Frontoparietal Control Network-A (FPN-A). See **Fig. 4A** and **4C** for example networks in one participant. These networks were selected for their relevance to MDD (e.g., Chen et al., 2020; Joormann and Vanderlind, 2014; Menon, 2011; Uddin, 2015), and individual networks were grouped based on topographical and functional properties. DN-A & DN-B are closely interdigitated networks implicated in internal mentation and together show “anticorrelation” to networks involved in external orienting (Du et al., 2024; see Buckner and DiNicola, 2019 for review). Within dlPFC, DN-A & DN-B tend to occupy more dorsal and medial regions. Moving ventrally, SAL & CG-OP are tightly interwoven networks that are linked to limbic reward circuitry and respond preferentially to salient targets (Du et al., 2024; Gordon et al., 2022; Seeley, 2019). Finally, FPN-A, which tends to be ventral to SAL & CG-OP, is implicated in cognitive control and recruited by cognitively demanding tasks in a domain-jlexible manner (Fedorenko, Duncan, and Kanwisher, 2013; DiNicola, Sun, and Buckner, 2023; Du et al., 2024). E-jield modeling of precision targeting (step 4) and network engagement characterization (steps 5 and 6) were performed for each target network set with the precision TMS pipeline.

### TMS Protocol

Repetitive TMS (rTMS) was administered to the modeled network sites in the second cohort of 8 participants as part of a broader study to assess the effects of stimulating distinct networks in participants with and without MDD. The broader study included administering rTMS to the left SAL & CG-OP, left FPN-A, and left DN-A & DN-B sites, with one site stimulated during 3 sessions in one day. All stimulation sessions used a neuronavigation system (Localite) to guide stimulation of the individualized targets (Localite GmbH, Bonn, Germany). First, the modeled coil orientation and position were inputted into Localite. Next, the participant’s anatomical landmarks were co-registered to their T1w image. The MagVenture Cool-B70 coil was then placed at the modeled site and secured with the MagVenture Flex Arm. When needed, minor adjustments were made to the coil position to maximize contact with scalp. In cases where the modeled coil orientation had the coil handle pointing anteriorly (towards the participant’s face), the orientation was flipped 180° and the current direction was reversed to achieve the equivalent E-field. rTMS was administered from the MagPro X100 stimulator with an intermittent theta burst protocol using the following parameters: 120% resting motor threshold (determined during the first MRI visit), 3 biphasic waveform pulses of 50Hz stimulation at a rate of 5Hz for a total of 1800 pulses per session, 8s inter-train interval, 570s total duration of each session. In cases of discomfort, the stimulation intensity was reduced to accommodate participants’ comfort on a case-by-case basis. Each target network was stimulated over 3 sessions, with each session separated by 45-50 minutes. Stimulation markers including the exact position of the coil center and orientation as well as the delivered dose were recorded by Localite. For each session, the average position, orientation, and dose were calculated and carried forth to E-field modeling using SimNIBS to retrospectively assess the reliability of the TMS coil placement and to estimate achieved network selectivity and intensity.

### Software and Statistical Analysis

Neuroimaging data storage and initial data quality assessment were conducted on XNAT (Marcus et al., 2007; https://www.xnat.org/). Preprocessing (iProc) used tools from FSL v5.0.4, FreeSurfer v6.0.0, and AFNI v16.3.13. Functional connectivity values were computed with MATLAB v2019a. The MS-HBM was implemented from: https://github.com/ThomasYeoLab/CBIG/tree/master/stable_projects/brain_parcellation/Kong2019_MS <u>HBM</u>. The TANS pipeline was implemented and adapted from: https://github.com/cjl2007/Targeted-Functional-Network-Stimulation. Head models and E-field simulations were calculated with SimNIBS v4.0.1. Visualizations of networks, correlation maps, target regions, and E-field maps in fsLR-32k space utilized Connectome Workbench v1.3.2. Selectivity and intensity plots were created using R ggplot2 (v4.2.2), and statistical analyses were conducted using R.

## Results

We first show how scalp landmark-based and sgACC anticorrelation-based TMS targeting influence distinct and multiple networks with variability between individuals. Then we present the TMS planning report from the automated, integrated pipeline that can be used to optimize coil positions to target specific networks within the brain of an individual. Finally, we present the results of applying this pipeline to target association networks that are near to one another within dlPFC to illustrate the potential of precision TMS targeting of networks, and further demonstrate that this targeting is feasible and reliable in a cohort of participants (including those with MDD) receiving real-world TMS with less than one hour of fMRI data acquired in a single session.

### Homotopic Scalp Landmark-Based dlPFC TMS Coil Placements Target Different Sets of Networks Within and Across Individuals

Left F3 and right F4 scalp positions are routinely used as dlPFC targets to treat MDD in TMS clinics. Precision mapping shows that the homotopic left F3 and right F4 TMS coil placements target different sets of networks to various extents within each participant. **Fig. 1A** shows network estimates in one example participant, and **Fig. 1B** shows the E-field from placing the coil at left F3 and right F4 with a 45° angle (relative to the sagittal plane). **Fig. 1C** quantifies the proportion of overlap between the E-field map, thresholded to the top 1% to 0.1%, for each network as a measure of spatial selectivity (Lynch et al., 2022). Further, as a measure of E-field intensity (**Fig. 1D**), the distribution of the E-field magnitude within each network is shown.

In this example participant, described in detail to make clear the challenges and opportunities of personalized modeling, no single network is selectively targeted. The top 0.5% of the left F3 E-field overlaps with the Salience (SAL) Network at 21.2%, and the Frontoparietal Control Network-B (FPN-B) at 8.4%. By contrast, at right F4, the top 0.5% of the E-field overlaps with SAL at 13.8% and FPN-B at 39.1%. The intensity plots suggest that the networks that received the highest stimulation intensity were the Frontoparietal Control Network-A (FPN-A) and the Cingulo-Opercular (CG-OP) Network at left F3, and FPN-A and FPN-B at right F4, with overall less high intensity stimulation on the right than the left. While the top 0.5% of the E-field at left F3 targeted SAL and the Default Network-B (DN-B) to similar extents as FPN-A and CG-OP, only FPN-A and CG-OP were exposed to the highest stimulation intensity. At right F4, FPN-B and FPN-A were the networks that overlapped the most with the top 0.5% of the E-field and were also the networks that received the highest stimulation intensity.

**Figure 1.**
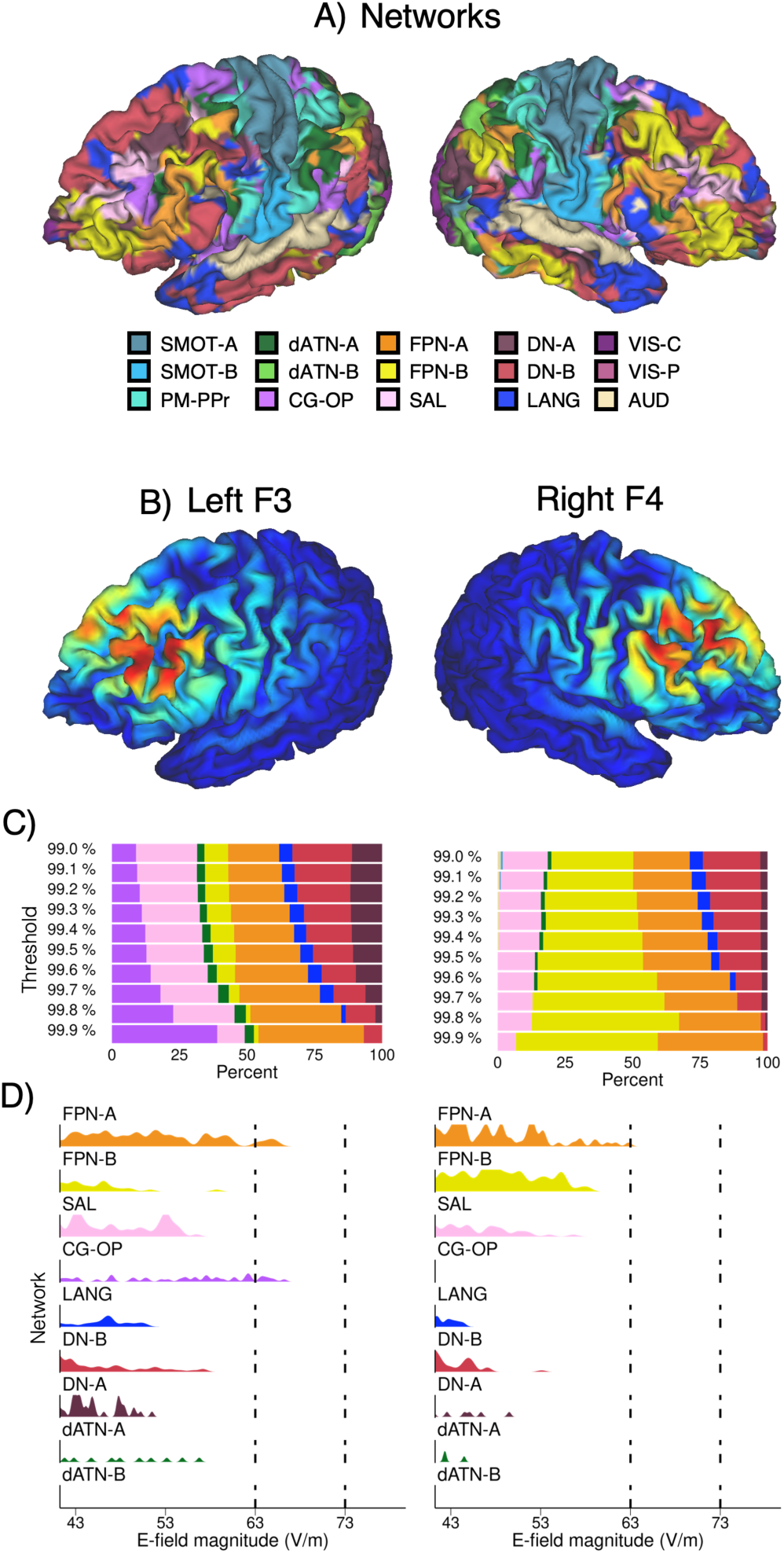
Left F3 versus right F4 TMS coil positions target distinct networks in a representative participant. Simulation results from one participant are shown with positioning of the TMS coil at left F3 and right F4 sites (homotopic scalp locations based on the Okamoto 10-20 EEG coordinate system). **A)** Network estimates on the left and right hemispheres of the cerebral cortex are presented. The network composition within the association zones varies between hemispheres including within dlPFC. The legend below labels the networks: SMOT-A, Somatomotor-A; SMOT-B, Somatomotor-B; PM-PPr, Premotor-Posterior Parietal Rostral; CG-OP, Cingulo-Opercular; SAL, Salience; dATN-A, Dorsal Attention-A; dATN-B, Dorsal Attention-B; FPN-A, Frontoparietal Network-A; FPN-B, Frontoparietal Network-B; DN-A, Default Network-A; DN-B, Default Network-B; LANG, Language; VIS-C, Visual Central; VIS-P, Visual Peripheral; AUD, Auditory. **B)** Maps of the E-Field effects are displayed for the left F3 and right F4 sites. Blue colors represent lower and red colors higher E-field (V/m) values. **C)** Overlap between the maps of the E-field model and network estimates quantified at various E-field thresholds are plotted. At left F3, the top 1% to 0.1% (99.0%-99.9%) E-field includes multiple networks without a clear predominant network. At right F4, there is relatively more FPN-B in the E-field, at each threshold. **D)** Distribution of E-field intensity values across the estimated networks with the E-field calibrated to dI/dt = 48 A/μS. Compared to other networks, there is higher stimulation intensity supplied to FPN-A and CG-OP on the left, and FPN-A and FPN-B on the right. The left hemisphere receives higher overall intensity than the right hemisphere.

On average across the 15 participants in the first cohort, at the top 0.5% of the E-field, left F3 most selectively targeted SAL (23.6%) and CG-OP (18.2%) and right F4 most selectively targeted SAL (26.3%) and FPN-B (22.1%). Notably, FPN-B was targeted with twice as much selectivity on the right (22.1%) versus the left (9.1%). **Fig. 2** shows that SAL is targeted similarly in terms of selectivity (at top 0.5% of the E-field) and maximal intensity (average of top 25 vertices) regardless of the hemisphere stimulated. However, FPN-B is generally targeted to a larger extent when the coil is positioned at right F4 compared to left F3.

**Figure 2.**
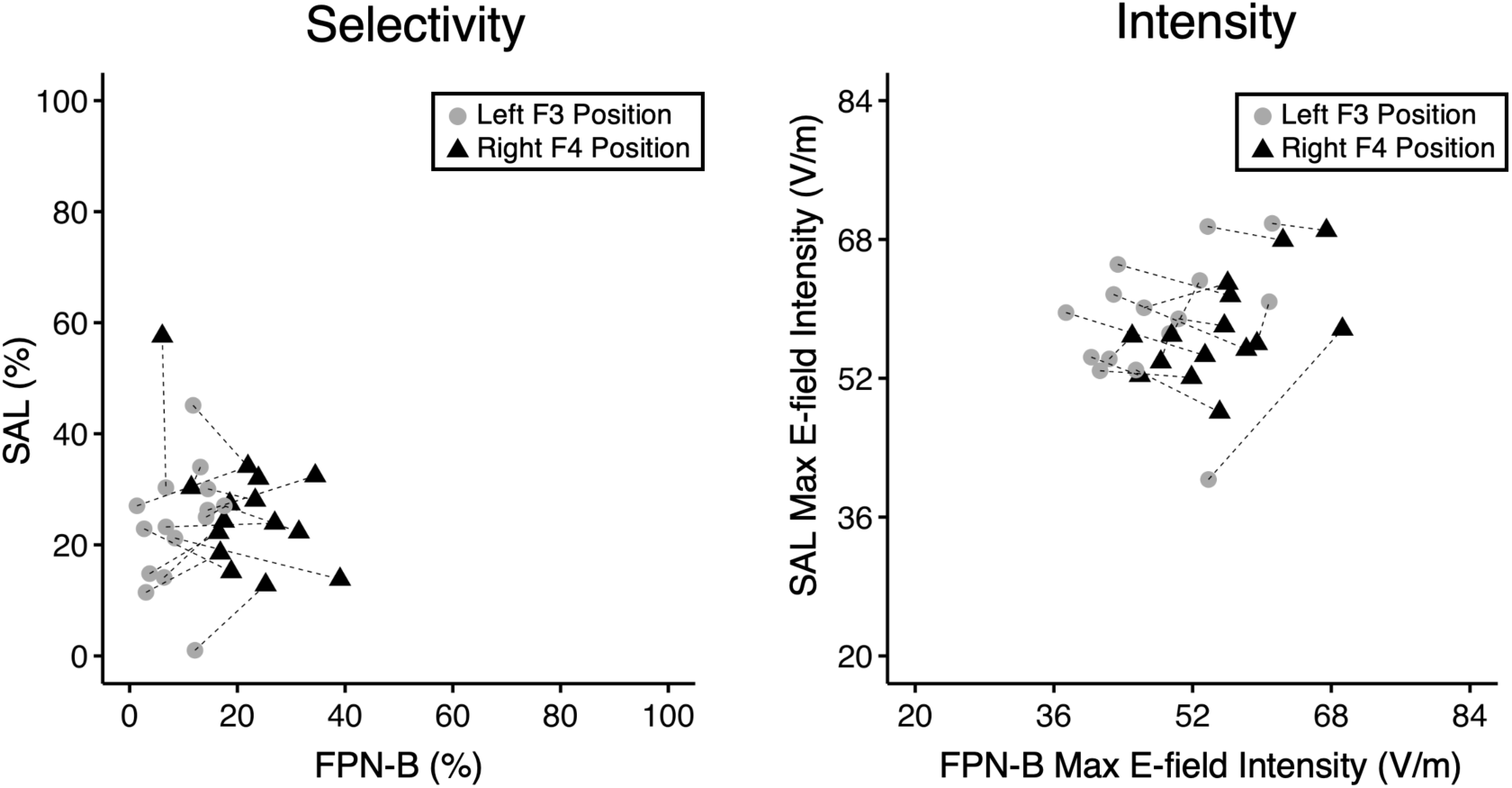
Selectivity and intensity for left F3 versus right F4 TMS coil positions quantified in 15 participants. Plots of selectivity and intensity illustrate the difference between the effects of scalp landmark-based left F3 and right F4 stimulation on the FPN-B versus SAL networks. Each participant is represented by one pair of connected symbols corresponding to the TMS coil placements: left F3, gray circle; right F4, black triangle. The left panel displays the selectivity corresponding to the relative % of SAL and FPN-B in the top 0.5% of the E-field. The right panel displays the intensity of the highest 25 vertices for FPN-B and SAL (each point is the mean of the highest 25 vertices). While the pairs of connected dots show an overall horizontal pattern reflecting that SAL is similarly targeted across hemispheres, the rightward shift for the right F4 estimates indicates that FPN-B, a right-lateralized candidate control network, is targeted more by positioning the TMS coil on the right hemisphere. Note also that the degree of SAL selectivity and intensity varies between participants considerably for both sites.

### The sgACC Anticorrelation Strategy Targets Different Networks Across Participants

Within-individual precision modeling revealed several insights about the zone of impact and potential therapeutic targets of the sgACC anticorrelation strategy. The sgACC correlation maps are consistent with previous findings showing that externally-oriented networks including Dorsal Attention-A (dATN-A) are anticorrelated to higher-order association networks involved in various aspects of internal mentation including DN-A & DN-B (Fox et al., 2005; Buckner and DiNicola, 2019). In general, within left dlPFC, CG-OP, SAL, and dATN-A were most strongly anticorrelated to the sgACC. FPN-A and LANG were neutral or modestly anticorrelated. On the other hand, DN-A, DN-B, and FPN-B were positively correlated to the sgACC (see **Fig. S1A**). That is, the correlated and anticorrelated regions (**Fig. S1B**) each contained representations of multiple distinct association networks.

**Fig. S2** demonstrates the sgACC anticorrelation strategy and networks in the anticorrelated target region for three example participants. The first row shows thresholded sgACC anticorrelations. The sgACC anticorrelated target region was defined as the largest contiguous anticorrelated patch on gyral crowns in dlPFC (**Fig. S2** second row). The networks falling within the anticorrelated target region were variable across participants, as shown in **Fig. 3A** and **Fig. S2** (third row). On average across participants, SAL composed 29.8% of this target region, with CG-OP at 32.7%, FPN-A at 18.6%, and dATN-A at 12.6% (**Fig. 3A**). In contrast, DN-A, DN-B, and FPN-B each composed <1.0% of the anticorrelated target region. Thus, while the sgACC anticorrelation approach avoids positively correlated networks (DN-A, DN-B, FPN-B), it does not consistently target a specific network in each individual.

The spatial selectivity and E-field intensity at the optimal coil placement for the anticorrelated target was examined. For all 15 participants in the first cohort, the network targeted with the highest selectivity (at top 0.5% of E-field) was determined: for 8/15, it was SAL (range: 25.9 – 52.2%), for 5/15, FPN-A (range: 23.0 – 49.8%), and for 2/15, CG-OP (range: 19.5 – 42.8%). Overall, across participants, the networks targeted with the highest selectivity were SAL (mean: 25.2%), FPN-A (mean: 23.0%), and CG-OP (mean: 18.8%). In addition, the network targeted with the highest intensity (average of top 25 vertices) was SAL for 6/15 participants (range: 54.3 – 69.7 V/m), FPN-A for 4/15 participants (range: 59.4 – 67.4 V/m), CG-OP for 3/15 participants (range: 59.3 – 81.2 V/m), and dATN-A for 2/15 participants (range: 62.0 – 65.0 V/m). Across participants, the networks targeted with the highest intensity were CG-OP (mean: 59.2 V/m), SAL (mean: 58.0 V/m), and FPN-A (mean: 57.5 V/m).

**Figure 3.**
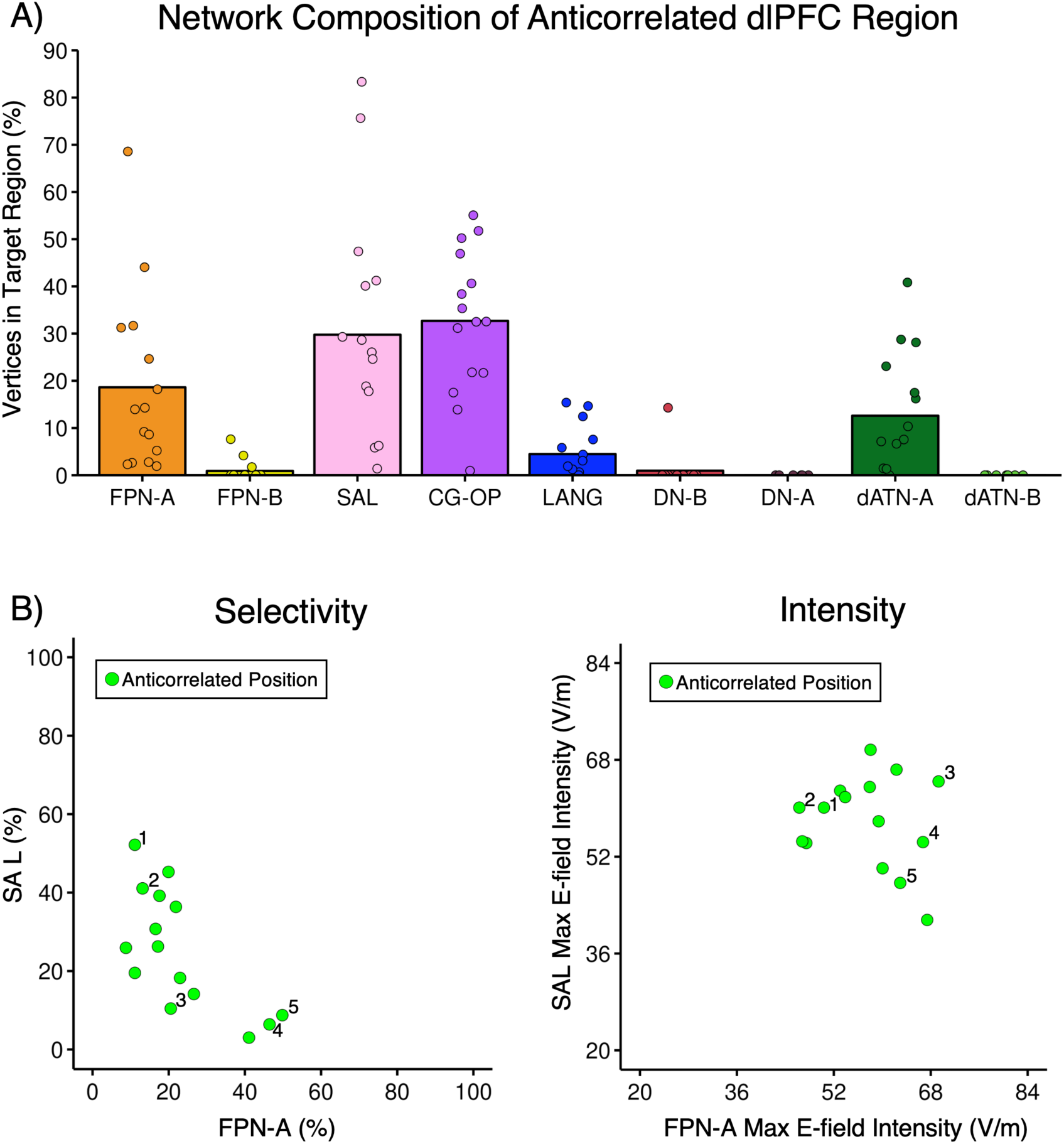
The sgACC anticorrelation strategy targets multiple networks with variability between individuals. Plots illustrate two facets of the sgACC anticorrelation targeting strategy. **A)** The region targeted by the sgACC anticorrelation contains subregions linked to multiple distinct networks. The plot displays the % of vertices in each network within the anticorrelated target region in 15 participants, with each bar representing the mean for a single network and the symbols representing individual participants. The sgACC anticorrelated target region variably includes SAL, CG-OP, FPN-A, and dATN-A network regions. Note specifically the marked variability between individuals in the degree to which the SAL network is included. **B)** Selectivity and intensity at the individualized sgACC anticorrelated target are quantified and plotted. Symbols represent the 15 individual participants. The left panel shows selectivity of the anticorrelated target, quantified as relative % of FPN-A versus SAL within the thresholded E-field (top 0.5% of values). The right panel shows the maximal intensity (mean of the 25 highest values) at the anticorrelated target for FPN-A versus SAL. The five highlighted participants (1-5) demonstrate that the degree of selectivity does not always predict the values for maximal intensity.

**Fig. 3B** shows, for all 15 participants in the first cohort, the relative proportion of SAL and FPN-A in the E-field thresholded at top 0.5% and the maximal intensity in these networks. Five participants are highlighted across the plots. In this sample, the degree of selectivity at the top 0.5% of the E-field does not always correspond with the maximal stimulation intensity received by each network. For example, highlighted participants 1 and 2 show selectivity for SAL (52.2% for participant 1 and 41.1% for participant 2) versus FPN-A (11.1% for participant 1 and 13.1% for participant 2), but a narrower difference in the maximal intensity received by each network (participant 1: SAL = 60.1 V/m, FPN-A = 50.4 V/m; participant 2: SAL = 60.1 V/m, FPN-A = 46.3 V/m). Participants 4 and 5 show the opposite network pattern but a similar contrast between selectivity and maximal intensity. These two participants demonstrate moderate selectivity for FPN-A and a narrower difference in maximal intensity received by FPN-A versus SAL & CG-OP. Participant 3 has low selectivity at the top 0.5% E-field (10.4% for SAL, 20.5% for FPN-A) and a relatively high maximal intensity in both networks (64.4 V/m for SAL, 69.3 V/m for FPN-A).

Overall, these plots suggest a higher separation between networks in terms of their exposure to the top 0.5% of the E-field (i.e., selectivity) but a lower separation in the maximal intensity received by each network, with some individuals showing a low degree of specificity across both measures.

### Automated Within-Individual Network Mapping and TMS Planning Report

To support precision network targeting in the clinic, we integrated individualized preprocessing, MS-HBM network estimation, TANS E-field optimization (Lynch et al., 2022), and characterization of network engagement into a cohesive pipeline. Critically, in this pipeline, pertinent results and visualizations are automatically compiled into a comprehensive TMS planning report. An example report is available in the Supplemental Materials (**Figs. S7-S12**). This specific report presents precision targeting results for left FPN-A, left SAL & CG-OP, left DN-A & DN-B, and right SAL & CG-OP, but can be adapted to other network targets or combinations of networks.

An adapted view of the first page of the report is displayed in **Fig. 4**. This page includes network parcellations, coordinates for the optimal coil position and orientation, target regions, E-field maps at the optimal coil placement, selectivity plots, and intensity plots (**Fig. S7**). The second page of the report includes scalp views showing selectivity for the target within the full search grid, and at each orientation for the optimal coil position (**Fig. S8**). This allows TMS operators to make adjustments due to real-world constraints such as participant comfort and coil contact with scalp. The third page of the report shows bilateral medial and lateral views of the E-field maps for each target, allowing for visualization of the spread of the E-field across cortex (**Fig. S9**). The fourth page shows how the target regions are selected on the inflated pial surface, with intermediate steps including search space masking, sulcal depth thresholding, and selection of the target network patch (**Fig. S10**). This allows for quality control of the target region as well as visualization of alternative targets should the primary region not be feasible or ideal. The fifth and sixth pages of the report display the optimal dose for each target network, assuming a neural activation threshold of 100 V/m (**Figs. S11-S12**).

### Distinct Juxtaposed Functional Networks Can be Targeted Within dlPFC Using Precision Modeling

Precision TMS targeting of three sets of these networks, interdigitated within the left dlPFC and thought to be involved in MDD, was initially tested in a cohort of 15 participants with extensive fMRI data collected over multiple sessions. The real-world applicability of this modeling approach was further demonstrated in an independent cohort of 8 participants with less than one hour of fMRI data acquired in a single session, including 4 participants with MDD. In both cohorts, the modeled selectivity of the stimulation for each target network (% On Target for the top 1% to 0.1% of the E-field) and the distribution of the intensity within each network were estimated. In the second cohort, which received TMS to the modeled network sites, the achieved selectivity and intensity were measured across multiple TMS sessions per target.

Precision targeting of the three network sets of interest is illustrated in **Fig. 4** in one example participant. After the networks were estimated for this participant (**Fig. 4A**), the largest contiguous cluster of the target network on the gyral crowns of the left dlPFC (entire ROI shown in **Fig. 4B**) was identified for each of the three target network sets (**Fig. 4C**). The E-field at the optimal coil placement for each target is displayed (**Fig. 4D**). The proportion of overlap between the E-field map thresholded at various levels (top 1% to 0.1%) and each network was then estimated as a measure of selectivity (**Fig. 4E**). The distribution of the E-field magnitude was quantified within each network as a measure of intensity (**Fig. 4F**).

The simulations in this participant showed a high degree of selectivity across the three network sets of interest. Each target network set comprised more of the top values of the E-field map relative to other networks, regardless of the threshold. The alignment between the top 0.5% of the E-field map and the target network was 49.5% for FPN-A, 79.8% for SAL & CG-OP, and 83.2% for DN-A & DN-B (**Fig. 4E**). Further, when considering intensity (**Fig. 4F**), the distribution of the E-field magnitude within each network shows that each target network set would receive the highest E-field strength among the higher-order networks. It is important to note that there is still uncertainty about the E-field magnitude required to activate neurons in different regions of association cortex. As a result, network-specific targeting needs to consider both selectivity and intensity. In this example, at the same stimulation dose, despite higher selectivity at the SAL & CG-OP and DN-A & DN-B optimized coil placements, network engagement may be higher at the FPN-A optimized coil position because its location exposes it to a higher intensity compared to the other two network sets.

**Figure 4.**
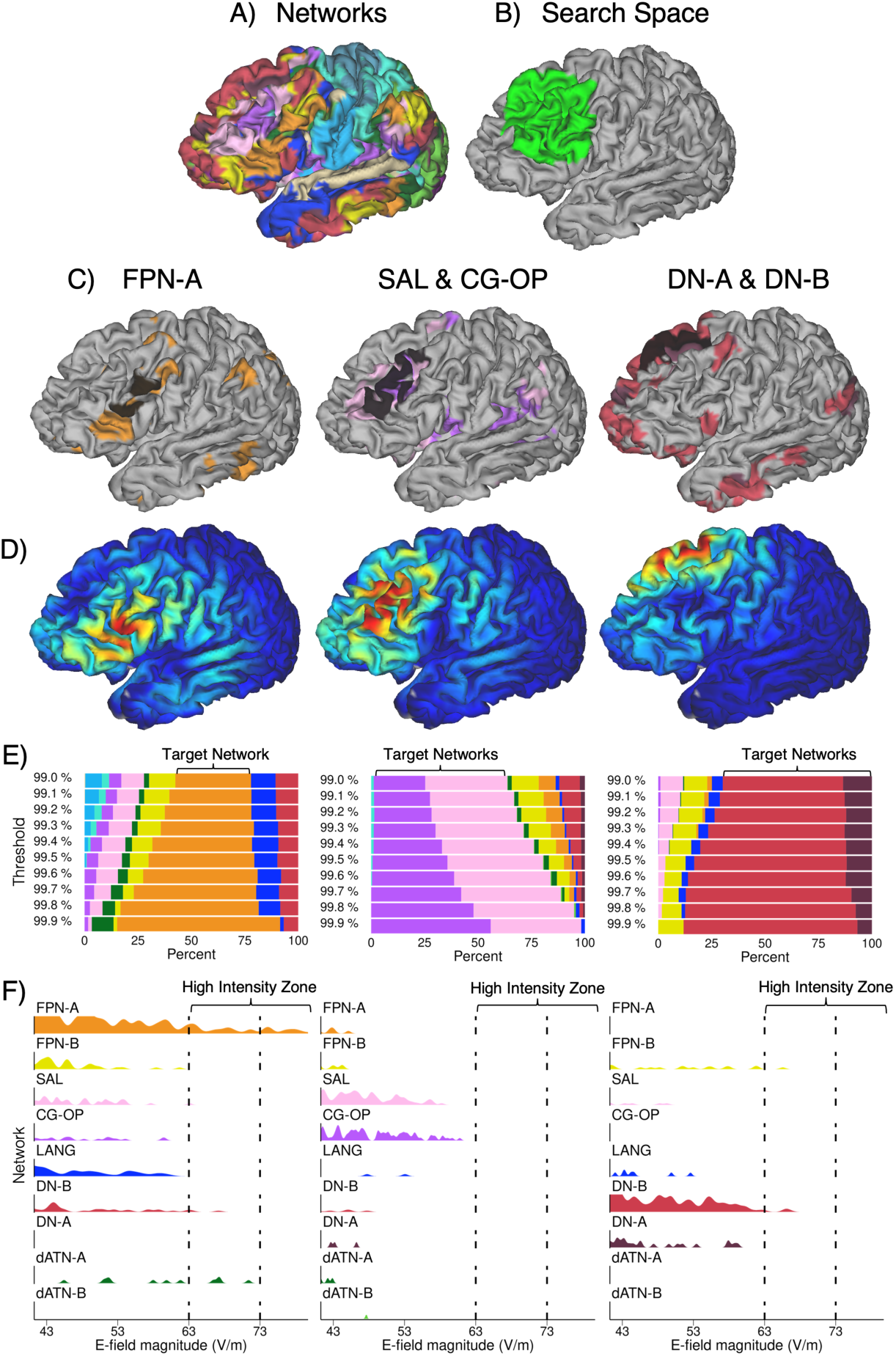
Precision network mapping and E-field modeling can be prospectively applied to target networks in individuals. Adapted views from the report generated by the precision TMS pipeline are displayed for one typical participant. **A)** Network estimates are displayed on the native-space surface with colors denoting distinct networks. The network colors are the same as in Figure 1. **B)** The dlPFC region used to constrain the search space for target selection is displayed in green. Note that the region overlaps with many distinct networks in panel A. **C)** The largest continuous cluster on a gyral crown within dlPFC is selected for each of the targets (highlighted in black) on top of the target network estimates shown in color. Note that an isolated network (e.g., FPN-A, left) or network pairs (e.g., DN-A & DN-B, right) can be set as targets, which illustrates the flexibility of this approach. **D)** The E-field map corresponding to the best coil placement for each target is shown. Blue colors represent lower and red colors higher E-field (V/m) values. **E)** The overlap between the E-field map and estimated networks is quantified at multiple thresholds (99.0%-99.9%). This participant demonstrates high spatial selectivity for each of the targets that increases as the thresholds increase. **F)** Plots show the distribution of E-field values within networks, with the E-field calibrated to dI/dt = 48 A/μS. Values on the right of the plot indicate high intensity stimulation, which is supplied to FPN-A in this participant, but not SAL & CG-OP or DN-A & DN-B. The E-field intensity plot considers all vertices in the cortex including those at depth and those outside the search space. It thus represents the estimated stimulation effect for the coil site and is independent of search space and target selection assumptions.

Among the first cohort of 15 intensively sampled participants, differential targeting of at least two distinct network sets was possible in most participants. The left column of **Fig. 5** shows the relative proportion of each pair of network sets in the E-field map thresholded at the top 0.5%. For each individually optimized coil placement, the top 0.5% of the E-field predominantly overlaps with the target network set. Specifically, the estimated degree of selectivity (i.e., % On Target) was >50% for 15/15 participants for the DN-A & DN-B networks, 8/15 for the SAL & CG-OP networks, and 4/15 for the FPN-A network (**Fig. 5 left**). The mean proportion of the E-field map thresholded at the top 0.5% overlapping with the target network was 76.0% (SD = 11.8%) for DN-A & DN-B, 53.2% (SD = 16.3%) for SAL & CG-OP, and 38.8% (SD = 15.1%) for FPN-A. The mean proportion of the E-field map thresholded at the top 0.5% overlapping with the non-target networks (e.g., FPN-A and DN-A & DN-B when the target is SAL & CG-OP) was 4.1% for DN-A & DN-B, 24.5% for SAL & CG-OP, and 22.1% for FPN-A.

While network-specific stimulation was estimated to be possible in most participants, various degrees of selectivity were found across network sets and participants. This suggests that variations in cortical geometry may lead to different capacities for precision TMS targeting. Notably, the degree of selectivity was significantly correlated with the target cluster size for FPN-A (*r* = 0.81, *p* < 0.001) and SAL & CG-OP (*r* = 0.87, *p* < 0.001), but not for DN-A & DN-B (*r* = 0.45, *p* = 0.09). This is likely the result of the FPN-A and SAL & CG-OP target clusters being smaller and more interdigitated than the corresponding DN-A & DN-B target clusters. These results thus indicate the necessity of large accessible network regions in the dlPFC for maximal selectivity in network targeting.

Further, motivated by findings from single-cell neurophysiology (Romero et al., 2019), which suggest that the effect of TMS stimulation is focal to the neurons receiving the highest intensity stimulation under the coil, we quantified the highest intensity delivered to target versus non-target network sets (**Fig. 5 right**). In this sample, at a pre-defined TMS dose of 35% MSO (dI/dt = 48 A/μS), when the coil placement was individually optimized to target DN-A & DN-B, the average maximal intensity (top 25 vertices) was 60.1 V/m for DN-A & DN-B, 29.0 V/m for FPN-A, and 36.7 V/m for SAL & CG-OP. These results indicate a clear separation between the target and non-target network sets in their simulated exposure to E-field energy. However, the differential intensity received was less when the coil was optimized to target the other two network sets of interest. When SAL & CG-OP was the target, the average maximal intensity (top 25 vertices) was 64.1 V/m for SAL & CG-OP, 48.5 V/m for FPN-A, and 48.2 V/m for DN-A & DN-B. When the coil position was optimized to target FPN-A, the maximal intensity was on average 67.3 V/m for FPN-A, 55.6 V/m for SAL & CG-OP, and 47.1 V/m for DN-A & DN-B. Therefore, while in most participants the three network sets could be targeted with relatively high selectivity (within the limits of TMS resolution), this targeting may still expose regions of the non-target networks to similar magnitudes of intensity in some participants.

**Figure 5.**
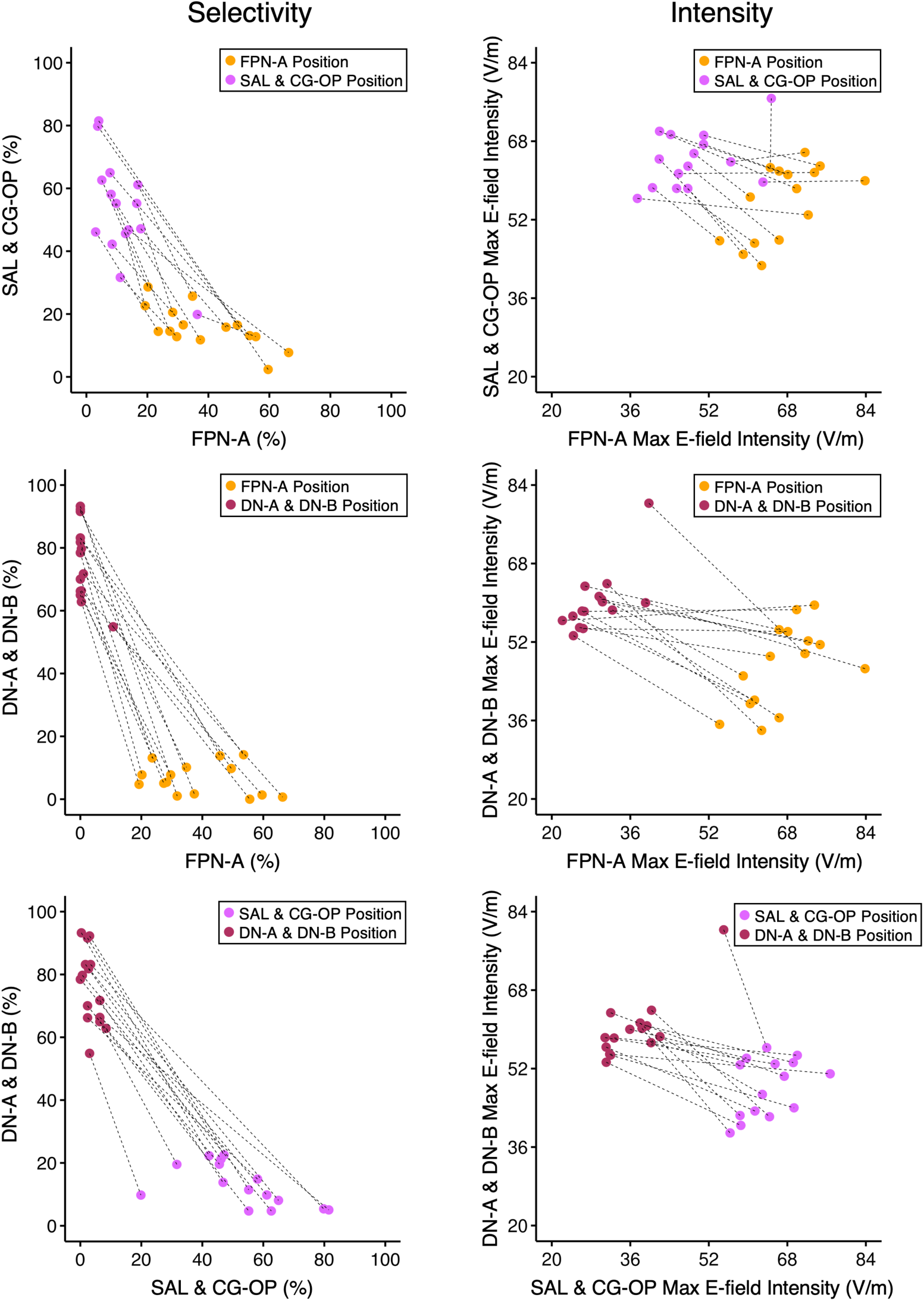
Hypothetical spatial selectivity and E-field intensity achievable in 15 participants. Plots display the modeled spatial selectivity and E-field intensity that is achieved by optimizing coil positions within individuals. Each pair of connected symbols represents one participant, and colors correspond to the optimal TMS coil placement for distinct networks: FPN-A, orange; SAL & CG-OP, purple; DN-A & DN-B, maroon. Left panels show the selectivity (relative % in the E-field at the top 0.5% of values); right panels show the maximal intensity of the highest 25 vertices (each point is the mean of the highest 25 vertices). The top row compares FPN-A versus SAL & CG-OP; the middle row compares FPN-A versus DN-A & DN-B, and the bottom row compares SAL & CG-OP versus DN-A & DN-B. Overall, there is clear separation in selectivity between the optimized coil positions, such that the coil placed at the target network(s) is more selective for the target than non-target network(s). This is also observed for maximal intensity, though the separation is narrower again reminding that the effects of selectivity and intensity can be distinct, and both should be taken into account.

Precision network estimation in the second cohort of 8 participants (N = 4 MDD) was feasible with less than one hour of fMRI data. Mirroring **Fig. 4**, precision targeting of the three network sets of interest is illustrated in **Fig. 6** in one example participant with MDD. In this participant, each individualized target demonstrated preferential selectivity for the intended networks (**Fig. 6E**). In addition, each target network set was modeled to receive the highest E-field intensity among the higher-order networks (**Fig. 6F**).

**Figure 6.**
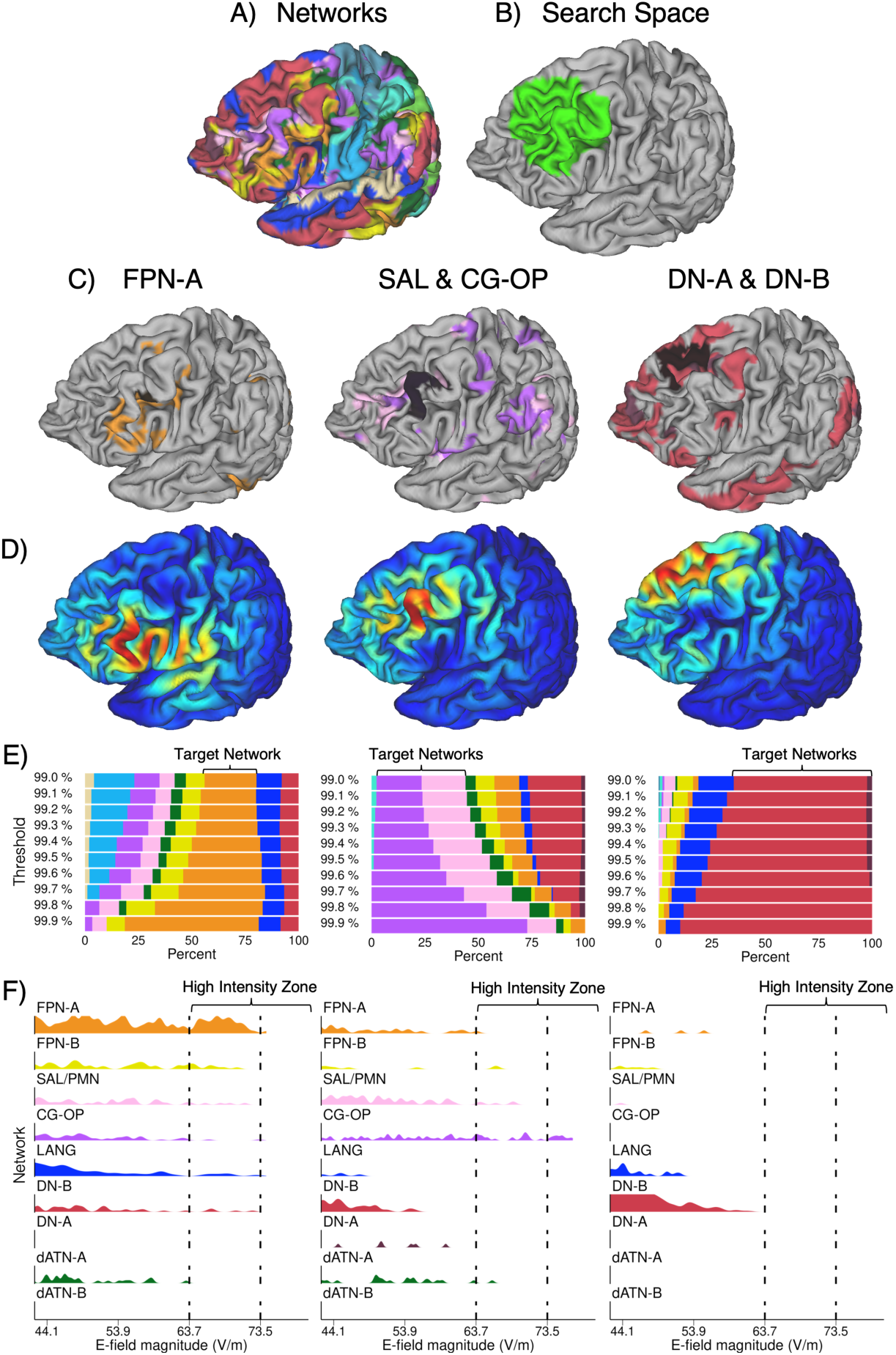
Precision network mapping and E-field modeling prospectively applied to target networks using single-session data in a participant with MDD. Mirroring the structure of Figure 4, using a single session of resting-state data (∼1 hr), the precision TMS pipeline was used to prospectively target distinct networks in a participant with MDD. **A)** Network estimates shown on the native-space surface. **B)** The dlPFC search space used for target selection. **C)** The network target regions (highlighted in black), overlayed on network estimates in color. **D)** The E-field map corresponding to the best coil placement for each target. Blue colors represent lower and red colors higher E-field (V/m) values. **E)** The overlap between the E-field map and estimated networks quantified at multiple E-field thresholds (99.0%-99.9%). This participant demonstrates preferential spatial selectivity for each of the targets that increases as the thresholds increase. **F)** Distribution of E-field values within networks, with the E-field calibrated to 120% of this participant’s resting motor threshold (dI/dt = 49 A/μS). Values on the right of the plot indicate high intensity stimulation, which is supplied to FPN-A and SAL & CG-OP in this participant, but not DN-A & DN-B.

Importantly, these modeled target sites were achievable during real-world TMS sessions, with preserved selectivity and intensity. Across all participants of the second cohort, separable degrees of selectivity were observed during actual TMS administration to modeled individualized targets (**Fig. 7 left**). This separation was consistent across multiple TMS sessions within each participant. These achieved results mirror the hypothetical estimates from the first cohort (**Fig. 5**), which underwent more intensive multi-session fMRI sampling. However, selectivity was lower in some individuals for FPN-A and SAL & CG-OP due to more interdigitated representations of these networks in dlPFC or discomfort during TMS at the prescribed targets. For the latter, coil placement was adjusted in the vicinity of the initial coil placement on a case-by-case basis to reduce discomfort while maintaining maximal network engagement, guided by the selectivity metrics of the automated report (**Fig. S8**).

Further, at 120% of each participant’s motor threshold, the maximal intensity (top 25 vertices) received by the target and non-target network sets demonstrated a clear separation in their estimated exposure to E-field energy during actual TMS sessions (**Fig. 7 right**). In addition, the achieved degree of network selectivity and intensity remained consistent across repeated TMS sessions, with intensity showing slightly higher inter-session variability than selectivity. This is illustrated in **Fig. 7**, where the symbols represent repeated sessions within individuals.

**Figure 7.**
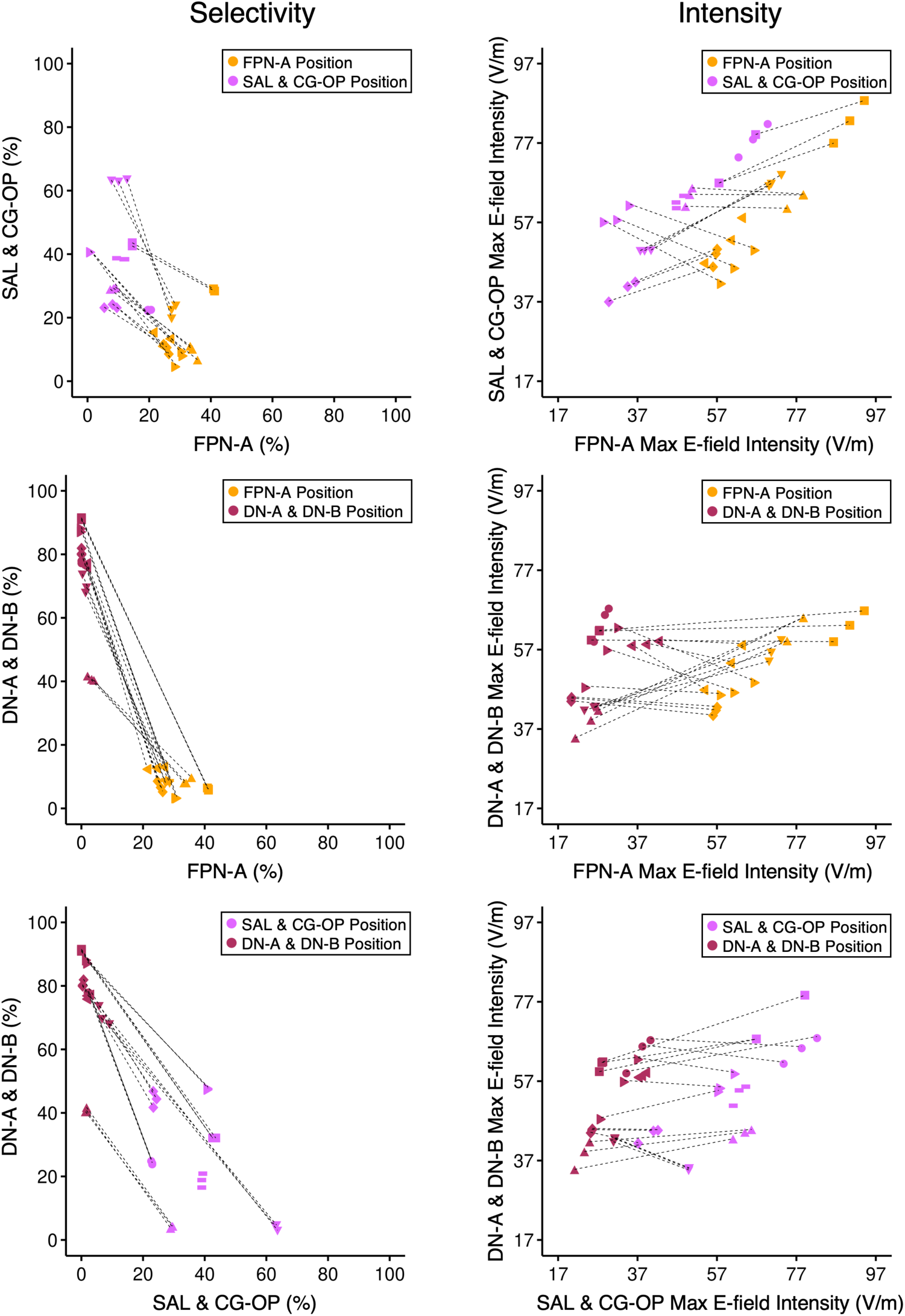
Achieved spatial selectivity and E-field intensity in 8 participants. Mirroring the structure of Figure 5, these plots display the achieved spatial selectivity and E-field intensity during real-world TMS sessions. Each symbol represents one participant, each pair of connected symbols represents the same TMS session number in the sequence of 3 TMS sessions administered per target, and colors correspond to the optimal TMS coil placement for distinct networks: FPN-A, orange; SAL & CG-OP, purple; DN-A & DN-B, maroon. Left panels show the selectivity (relative % in the E-field at the top 0.5% of values); right panels show the intensity of the highest 25 vertices (each point is the mean of the highest 25 vertices). Overall, there is clear separation in selectivity and maximal intensity between the optimized coil positions, such that the coil placed at the target network(s) is more selective and achieves higher intensity for the target than non-target network(s).

## Discussion

The present study advances our understanding of TMS targeting by combining individualized precision network mapping and E-field modeling. Our findings first characterize potential therapeutic targets in current clinical TMS protocols at the network level within individuals. We then demonstrate the feasibility and reliability of using precision TMS targeting to preferentially engage distinct functional networks within the dlPFC, even when the networks are near to one another and variable between individuals. Precision TMS targeting can potentially be applied in the clinic to assess both the prospective and the retrospective functional network engagement of TMS targets within individuals, which can be used to guide decision-making and TMS efficacy assessment.

### Within- and Between-Individual Variability in Standard Clinical Scalp Landmark-Based TMS

The present within-individual analyses converge with prior group-based network estimates to show that multiple networks are engaged by using standard clinical targets (Harita et al., 2022; Cardenas et al., 2022). Our results extend beyond these analyses of average target engagement to reveal the variability between individuals and hemispheres. For example, several clinics employ left versus right scalp-based targets depending on whether a patient’s presenting symptoms are predominantly anhedonic or anxiosomatic, respectively (Cirillo et al., 2019; Diefenbach et al., 2016; Siddiqi et al., 2020; Zwanzger et al., 2009). Our analysis of two standard homotopic scalp landmark-based targets (left F3 and right F4) revealed considerable variability in the networks engaged within and across individuals. There was no clear dominant network targeted by F3 in the left hemisphere nor by F4 in the right hemisphere, but some patterns emerged.

First, at both sites the SAL network, a network linked to reward circuitry, had the highest spatial selectivity at the top 0.5% of the E-field, albeit modestly. Second, at right F4, FPN-B, a right-lateralized association network which tends to be positively correlated with the sgACC (see **Fig. S1**), is targeted to a greater extent than at left F3, in terms of both selectivity and intensity. While the specific function of FPN-B remains elusive, its position suggests a role in cognitive control possibly aligned to affective domains (Du et al., 2024). It is thus of interest that preferential targeting of FPN-B in the right hemisphere is the main difference we observed between F3 and F4 coil positions and warrants further investigation. Therefore, the variability in network engagement incurred by standard clinical TMS based on scalp landmarks across hemispheric allocations may contribute to the heterogeneous clinical responses observed in TMS treatment.

### Networks Impacted by the sgACC Anticorrelation Strategy

The sgACC anticorrelation strategy showed variability in the networks targeted across participants. While this approach consistently avoided DN-A, DN-B, and FPN-B, the network preferentially targeted and receiving the most intense stimulation varied between SAL and FPN-A across individuals. When considering both selectivity and intensity, SAL appeared to be the dominant network targeted by the anticorrelation approach. This is a particularly interesting finding in light of evidence that the SAL network (1) is coupled to regions in the ventral striatum consistent with a role in reward processing (Gordon et al., 2022; Seeley, 2019), (2) may be topographically expanded in depressed individuals as demonstrated by precision MRI mapping (Lynch et al., 2024), and (3) is comprised of regions (e.g., the anterior and ventral insula) densely anatomically interconnected to circuits implicated in autonomic and interoceptive processing (e.g., Benarroch, 2019; Evrard, 2019; Seeley, 2019; Craig, 2002; Fischer et al., 2016). Our findings raise the possibility that the antidepressant mechanism of anticorrelation-based strategies like SNT is the stimulation of specific canonical networks such as the Salience network—rather than a more generalized modulation of anticorrelated dlPFC regions. Future studies could empirically test this hypothesis by using the present (or similar) methodology to examine the relations between clinical efficacy and the network (or networks) targeted.

### Differential Precision Targeting of Juxtaposed Functional Networks in the Individual

Our results show that it is possible to differentially target functional networks that are closely juxtaposed within dlPFC with high selectivity and intensity in the individual. This precision may be important given the role different networks serve in different neuropsychiatric symptom profiles (e.g., Williams, 2016) and given that, in practice, many neuropsychiatric disorders are clinically heterogeneous (Drysdale et al., 2017). For example, in patients with internally oriented ruminative symptoms, it may be beneficial to target DN-A & DN-B with TMS. By contrast, in patients with symptoms related to threat dysregulation and anxiety, SAL & CG-OP could be targeted. Similarly, patients experiencing symptoms related to deficits in cognitive control, such as emotion dysregulation, may benefit from TMS targeting FPN-A. Indeed, prior work suggests that TMS can induce network-specific changes in functional connectivity and metabolism (Eldaief et al., 2011; 2023). Taken together, our targeting approach could be used to preferentially stimulate different networks in dlPFC in different patients depending on their presenting symptom profiles. The clinical utility of symptom-to-target matching is unknown at this time, and the main driver of efficacy may be more predicted by effective dose than network-specific targeting. The present methods provide a way to test specific alternative targets in head-to-head comparisons. Alternatively, to the degree clinical outcomes are available along with fMRI and recorded coil positions, it may be possible to correlate outcomes and targets to gain insights into what matters and what is possible.

It is important to note that individual anatomy and cortical geometry may limit the ability to selectively target certain networks in certain individuals. We found that target size is an important factor, with larger target regions for FPN-A and SAL & CG-OP predicting higher degrees of selectivity. As such, our methodology may be less useful in individuals who have smaller representations of relevant networks in dlPFC. Relatedly, an individual’s network organization in dlPFC may be such that certain networks are preferentially represented deeper within the fundus of a sulcus and therefore less accessible to TMS. Importantly, in these cases our precision network modeling approach could ostensibly be used to target network representations outside of dlPFC but nevertheless anatomically situated closer to the scalp (e.g., portions of parietal cortex or the cerebellum).

### Importance of Spatial Selectivity and E-field Intensity in Precision TMS

The present approach allows for the identification of optimal coil placements and estimation of two measures to assess the effect of network-specific targeting: spatial selectivity and E-field intensity. Selectivity, or the proportion of overlap between the target network and the top values of the E-field, is the primary measure used by TANS to optimize coil placement (Lynch et al., 2022). While this measure is critically important, it relies on an arbitrary cutoff looking at the top percentage of values without considering the maximal intensity received in different parts of the cortex (Numssen et al., 2024). Neurophysiological work that combines TMS with invasive intracranial recordings suggests that the effects of TMS may be more focal than E-field spread suggests (Romero et al., 2019) and neurons receiving a higher E-field are more likely to exhibit intracranial TMS evoked potentials (Wang et al., 2024). Thus, it is important to also consider the maximal and overall intensity of the E-field supplied to target networks. Our results show that high selectivity does not necessarily correspond to maximal intensity (e.g., **Fig. 3B**). Further experiments are needed to discern how these factors predict TMS efficacy, and toward this end, we provide a tool to assess both network-level selectivity and intensity.

### Limitations and Future Directions

While our study demonstrates the potential for precision network TMS modeling, several limitations should be noted. As mentioned above, the importance of selectivity versus intensity in TMS targeting remains unclear and cannot be addressed with our present methodology. For instance, clinical efficacy could ostensibly be achieved by stimulating the network mediating clinical benefits with sufficient intensity, even if adjacent but less clinically relevant networks are also stimulated. Alternatively, unwanted off-target network effects could limit efficacy, mandating a high level of network selectivity. As another possibility, the effective dose may be different between sites with maximal stimulation being the primary driver of efficacy without a relation to network composition. Our modeling approach could be used to resolve these questions by relating clinical outcomes to the degree of selectivity versus maximal intensity achieved.

Active translational efforts in our lab are focused on modeling and modulating functional networks in individuals with treatment-resistant depression. Results presented in this study suggest that precision network TMS is feasible in such patients and under conditions where a single one-hour session of functional data is acquired to estimate networks (rather than the multi-session acquisitions utilized in the first cohort of this study). In addition, validation experiments will be needed to demonstrate that stimulating distinct networks yields distinct behavioral and clinical outcomes. The present precision TMS pipeline is constructed to assist studies that seek to test the effects of network-specific TMS on different cognitive domains and symptoms.

Another limitation arises from our use of E-field models. The results from this study are model estimates and are thus limited by the accuracy of the parameters used in the computational models, and the general assumption that the models appropriately capture all relevant biophysical properties. These parameters include the TMS coil model, tissue segmentation and estimated conductivity. More subtle aspects such as anisotropy of the conductivity in the white matter and meninges (Weise et al., 2022), for example, are not considered.

When executing precision TMS in the real world, several practical factors must also be considered. Given the complexity of the pipeline, checks were performed to ensure adequate data quality, registration, and model quality. These checks are enabled by the QC outputs of the precision TMS pipeline and include visualization of the registration of structural and functional images, which is necessarily imperfect given the distortion inherent to functional neuroimaging. In addition, precise coil positioning may be affected by discomfort, physical constraints to optimize contact between the coil and the scalp, or head movements during longer TMS sessions. Further, robotic TMS systems may offer an interesting solution to improve precision targeting accuracy. These practical considerations will impact the degree to which model estimates of TMS stimulation effects will translate to the actual practice of clinical TMS.

### Conclusions

We characterize the within-individual network engagement of current clinical TMS strategies, present an integrated pipeline for precision network TMS targeting, and demonstrate the feasibility and reliability of differentially targeting specific networks with meaningful selectivity and intensity. This work provides a path towards relating clinical outcomes to individual-specijic, systems-neuroscience informed functional anatomy.

## Data Availability

All data produced in the present study are available upon reasonable request to the authors.

## Acknowledgements

We thank the Harvard Center for Brain Science neuroimaging core and FAS Division of Research Computing. We thank Tim O’Keefe and Daniel Asay for assisting in optimization of data processing, Ross Mair for MRI physics support, Kathryn Rodrigues for assisting with proofreading, Joanna Ladoupoulou, Vaibhav Tripathi, Max Elliott, Heather Kosakowski, Oula Puonti, and Nolan Williams for helpful feedback and discussion, and Conor Liston’s group for making the Targeted Functional Network Stimulation (TANS) code and protocol openly available.

